# A statistical genomics framework to trace bacterial genomic predictors of clinical outcomes in *Staphylococcus aureus* bacteraemia

**DOI:** 10.1101/2022.04.21.22273941

**Authors:** Stefano G. Giulieri, Romain Guérillot, Natasha E. Holmes, Sarah L. Baines, Abderrahman Hachani, Diane S. Daniel, Torsten Seemann, Joshua S. Davis, Sebastiaan Van Hal, Steve Y. C. Tong, Timothy P. Stinear, Benjamin P. Howden

## Abstract

Outcomes for patients with severe bacterial infections are determined by the interplay between host, pathogen, and treatments. Most notably, patient age and antibiotic resistance contributes significantly to poor outcomes. While human genomics studies have provided insights into the host genetic factors impacting outcomes of *Staphylococcus aureus* infections, comparatively little is known about *S. aureus* genotypes and disease severity. Building on the idea that bacterial pathoadaptation is a key driver of clinical outcomes, we develop a new genome-wide association study (GWAS) framework to identify adaptive bacterial mutations associated with clinical treatment failure and mortality in three large and independent *S. aureus* bacteraemia cohorts, comprising 1358 episodes. We discovered *S. aureus* loci with previously undescribed convergent mutations linked to both poorer infection outcomes and reduced susceptibility to vancomycin. Our research highlights the potential of vancomycin-selected mutations and vancomycin MIC as key explanatory variables to predict SAB severity. The contribution of bacterial variation was much lower for clinical outcomes (heritability < 5%), however, GWAS allowed us to identify additional, MIC-independent candidate pathogenesis loci. Using supervised machine-learning, we were able to quantify the predictive potential of these adaptive *S. aureus* signatures, along with host determinants of bacteraemia outcomes. The statistical genomics framework we have developed is a powerful means to capture adaptive mutations and find bacterial factors that influence and predict severe infections. Our findings underscore the importance of systematically collected, rich clinical and microbiological data to understand bacterial mechanisms promoting treatment failure.

## INTRODUCTION

The course and outcome of severe bacterial infections such as bacteraemia and sepsis are determined by the interaction of factors related to the host (e.g. genetics, age, chronic conditions), the pathogen and the treatment (Cohen, Vincent et al. 2015). A deeper understanding of these outcome determinants would help design more effective treatments and inform precision medicine strategies. However, in contrast with progress in the immunogenetics of infections (Kwok, Mentzer et al. 2021), it has been challenging to identify bacterial genetic factors influencing infection outcomes (Giulieri, Tong et al. 2020, Denamur, Condamine et al. 2022). This is due to the considerable genetic diversity of bacterial pathogens and the strong clonal structure of bacterial populations that complicate genome-wide association studies (GWAS) (Power, Parkhill et al. 2017).

With up to 20% mortality (Bai, Lo et al., Battle, Shuping et al. 2021) and increasing incidence (Imam, Tempone et al. 2019), *Staphylococcus aureus* bloodstream infections are a key challenge to human health (Tong, Davis et al. 2015). The clinical determinants of *Staphylococcus aureus* bacteraemia (SAB) outcomes have been described extensively, in particular mortality has been the subject of numerous studies (reviewed here (van Hal, Jensen et al. 2012)). Small studies have shown the impact of host genetic factors of SAB risk and outcomes, for example it was shown that a polymorphism in the DNA methyltransferase *DNMT3A* protects from persistent SAB (Mba Medie, Sharma-Kuinkel et al. 2019). Studies linking specific clones or bacterial characteristics (vancomycin minimum inhibitory concentration [MIC]) with SAB mortality (Holmes, Turnidge et al. 2011) provide indirect evidence for the role of bacterial genetic factors in SAB mortality, however, it has been difficult to identify compelling associations between distinctive genes or mutations and clinical outcomes (Giulieri, Holmes et al. 2016). In a landmark study Recker *et al*, used a combination of GWAS and machine learning to explore factors associated with SAB mortality. Their work showed that both bacterial genetic and phenotypic information improved the performance of the predictive model. Feature selection from the model indicated several potential pathogenic mutations, that could represent the basis for functional validation (Recker, Laabei et al. 2017).

A key concept underlying bacterial phenotype-genotype studies is pathoadaptation, where bacteria acquire mutations that enhance their ability to survive in the host. Upon infection, colonising *S. aureus* strains are repeatedly subjected to similar selective pressures; thus, mutations promoting survival in the host during infection are expected to be preferentially maintained and become evident when examining *S. aureus* microevolution. The most compelling evidence for the role of pathoadaptation in SAB comes from within-host evolution studies, which have shown enrichment of mutations in a small number of genes including *agr* and antibiotic resistance genes (indicating convergent evolution), both upon transition from colonisation to invasion and during persistent infection (Young, Wu et al. 2017, Giulieri, Baines et al. 2018). Bacterial GWAS leverage the concept of adaptation either directly by focusing on homoplastic mutations (phylogeny-based methods) (Farhat, Shapiro et al. 2013, Brynildsrud, Bohlin et al. 2016, Collins and Didelot 2018, Saund and Snitkin 2020, Chen and Shapiro 2021) or indirectly by correcting the analysis for population structure (Purcell, Neale et al. 2007, Zhou and Stephens 2012, Earle, Wu et al. 2016, Lees, Galardini et al. 2018). For example, Farhat *et al* identified multiple resistance-associated mutations and genes by applying a phylogenetic convergence testing approach to a collection of *M. tuberculosis* isolates (Farhat, Shapiro et al. 2013). Convergent evolution was also used in GWAS of *M. tuberculosis* tolerance (Hicks, Yang et al. 2018), and vancomycin-intermediate *S. aureus* (VISA) phenotype (Alam, Petit et al. 2014). Approaches based on population structure (allele-counting methods) don’t account for convergent evolution explicitly, but the most robust associations are expected to occur with homoplastic mutations or genes under positive selection. For example, in a GWAS of drug resistance in *M. tuberculosis*, 50% of gene targets identified using regression and correction for population structure were shown to be homoplastic using a convergence-based approach (Farhat, Freschi et al. 2019).

The examples described above involve antibiotic resistance, an adaptive phenotype with high heritability and strong selective pressure, where convergent evolution is expected to be apparent. However, it is unclear whether clinical outcomes of bacterial infections are dependent on bacterial genotypes, as shown by bacterial GWAS of clinical outcomes, that have been either negative or have not been reproduced across independent cohorts. Bacterial adaptation can be frequently detected in clinical infections (Culyba and Van Tyne 2021), leading to antibiotic resistance, antibiotic tolerance, or immune evasion, and thus potentially driving treatment failure (Balaban, Helaine et al. 2019). Here, we propose a statistical genomics framework that incorporates convergent evolution explicitly and tracks molecular signatures of adaptation across three independent SAB patient cohorts to build a model of bacterial genotype-phenotype associations for treatment outcomes. Given the complexity of identifying genetic correlates of clinical outcomes, we progress our analysis from outcomes with high expected role of adaptation (vancomycin MIC) to intermediate outcomes (duration of bacteraemia) to final endpoints with strong multi-factorial pathogenesis (treatment success, mortality).

## RESULTS

### *S. aureus* bacteraemia cohorts

To identify bacterial genetic correlates of clinical outcomes of bacteraemia, we analysed 1,358 *S. aureus* sequences from 3 cohorts of SAB with extensive clinical metadata. The cohorts included an observational study of vancomycin efficacy in SAB (cohort A, 843 isolates, 738 individual episodes), an observational study of the VISA/heterogenous VISA phenotype in sequence type (ST) 239 methicillin resistant *S. aureus* (MRSA) (cohort B, 296 isolates) and a randomised clinical trial of vancomycin-β-lactam combination therapy in MRSA bacteraemia (cohort C, 324 isolates). The microbiological and clinical characteristics of the isolates are described in Table 1 and Figure 1. The cohorts were different in terms of proportion of MRSA, distribution of sequence-types, mortality outcomes and treatment choice. Therefore, rather than combining all sequences in a single dataset, we analysed the three collections separately, with the aim of finding overlapping hits between the independent samples that would be provide robust evidence for clinically relevant bacterial adaptation.

**Table 1.** Characteristics of *S. aureus* isolates included in the bacteraemia cohorts

|  | **Cohort A**<br>**(n = 738)** | **Cohort B**<br>**(n = 296)** | **Cohort C**<br>**(n = 324)** |
| --- | --- | --- | --- |
| Sample period |  |  |  |
| 1990-1999 | 0 | 32 (11) | 0 |
| 2000-2009 | 517 (70) | 264 (89) | 0 |
| 2010-2019 | 221 (30) | 0 | 324 (100) |
| Country |  |  |  |
| Australia/NZ | 738 (100) | 296 (100) | 239 (74) |
| Singapore | 0 | 0 | 53 (16) |
| Israel | 0 | 0 | 32 (10) |
| Patient age (years), median (IQR) | 61 (46-74) | 64 (50-73) | 64 (49-76) |
| MRSA | 252 (34) | 296 | 324 |
| Vancomycin MIC (mg/L), median (IQR) | 1.5 (1-2) | 1 (0.75-1.5) | N/A |
| Main antibiotic |  |  |  |
| Flucloxacillin | 384 (52) | 0 | 0 |
| Vancomycin | 301 (41) | 252 (85) | 158 (49) |
| Daptomycin | 0 | 0 | 3 (1) |
| Vancomycin +β-lactam | 0 | 0 | 159 (49) |
| Daptomycin+ β-lactam | 0 | 0 | 4 (1) |
| Other | 53 (7) | 11 (4) | 0 |
| No treatment | 0 | 28 (9) | 0 |
| Duration of bacteraemia (days), median (IQR) | 1 (1-2) | N/A | 1 (1-3) |
| Mortality at 30 days | 109 (15) | 88 (30) | 35 (11) |

Values are counts (percent) unless otherwise specified.
MRSA: methicillin-resistant *S. aureus*. N/A: not available.

**Figure 1.**
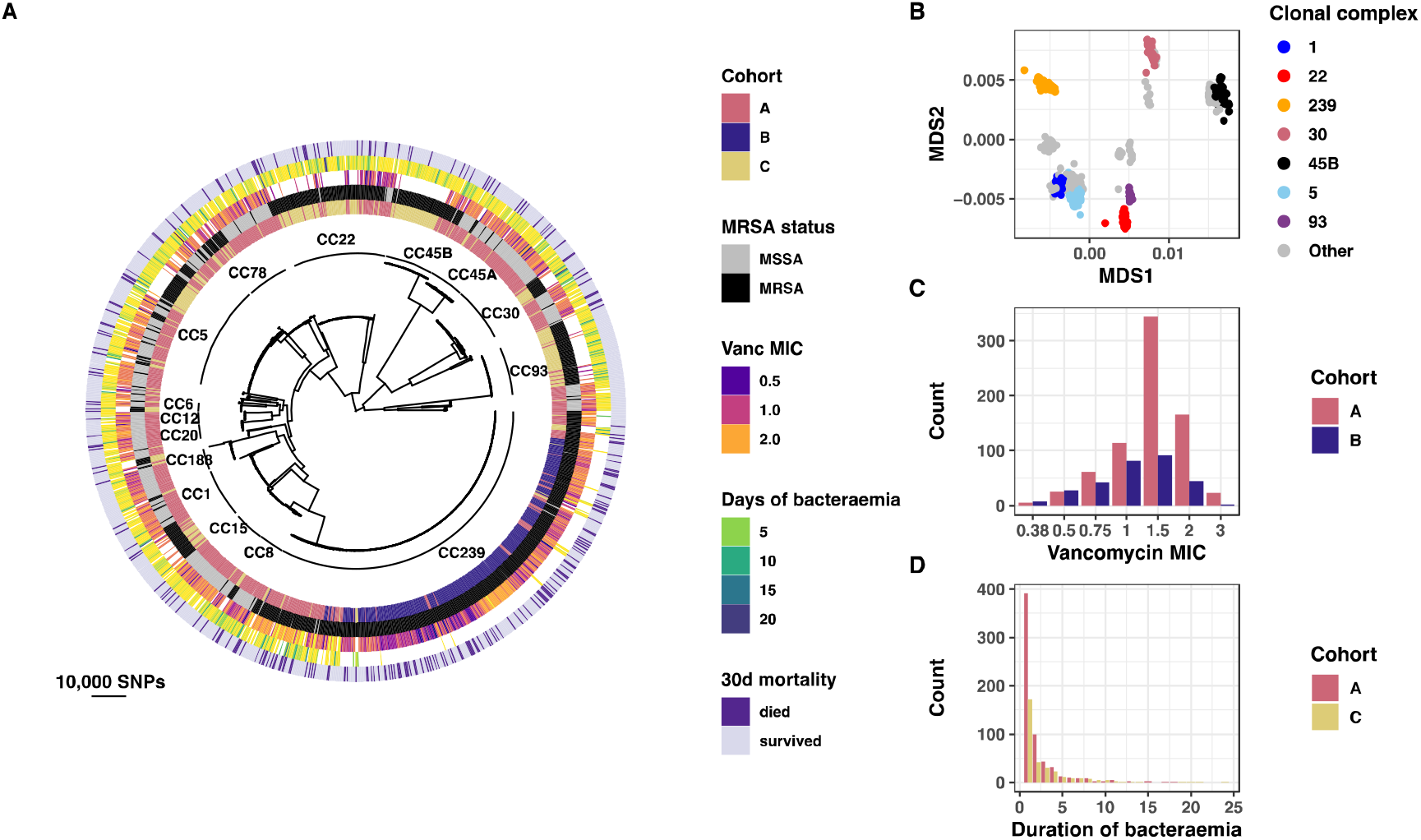
Genomic and phenotypic characterisation of 3 *S. aureus* bacteraemia cohorts. (A) Maximum likelihood phylogenetic trees of 1,358 bloodstream *S. aureus* strains included in the analysis. (B) First two principal components of multi-dimensional scaling (MDS) of the Mash distance matrix. Dots represent sequences and are coloured according to the 7 most prevalent clonal complexes (C) Distribution of vancomycin MIC (in mg/L). (D) Distribution of duration of bacteraemia (in days).

### Lineage effects and bacterial adaptation may impact clinical outcomes of *S. aureus* bacteraemia

We first assessed the relative impact of clinical factors on SAB outcomes using cohort A because it included all three outcomes of interest for this study (vancomycin MIC, duration of bacteraemia, mortality) (543 episodes) and then fitted a classification model of mortality. In our initial model we included predictors of SAB mortality that are generally available in the clinic including host factors (age, gender, comorbidities, site of acquisition), anti-staphylococcal treatment, phenotypic bacterial characteristics (methicillin-resistance, vancomycin MIC) and duration of bacteraemia.

We used a random forest machine learning algorithm that was trained using stratified cross-validation on 80% of cohort A episodes. To increase the accuracy of the predictive parameters estimates the model was trained on 100 replicate training-testing data splits and produced a mean area under the receiver-operating curve (AUROC) of 0.803 and mean area under the precision-recall curve of 0.395 (AUPRC) (Figure 2A). Thus, traditional predictors of mortality in SAB had an acceptable predictive performance (Lemeshow, Sturdivant et al. 2013), however, 20% of mortality predictions were incorrect, raising the possibility that the addition of non-clinical variables (including bacterial genomic features) could increase prediction accuracy.

**Figure 2.**
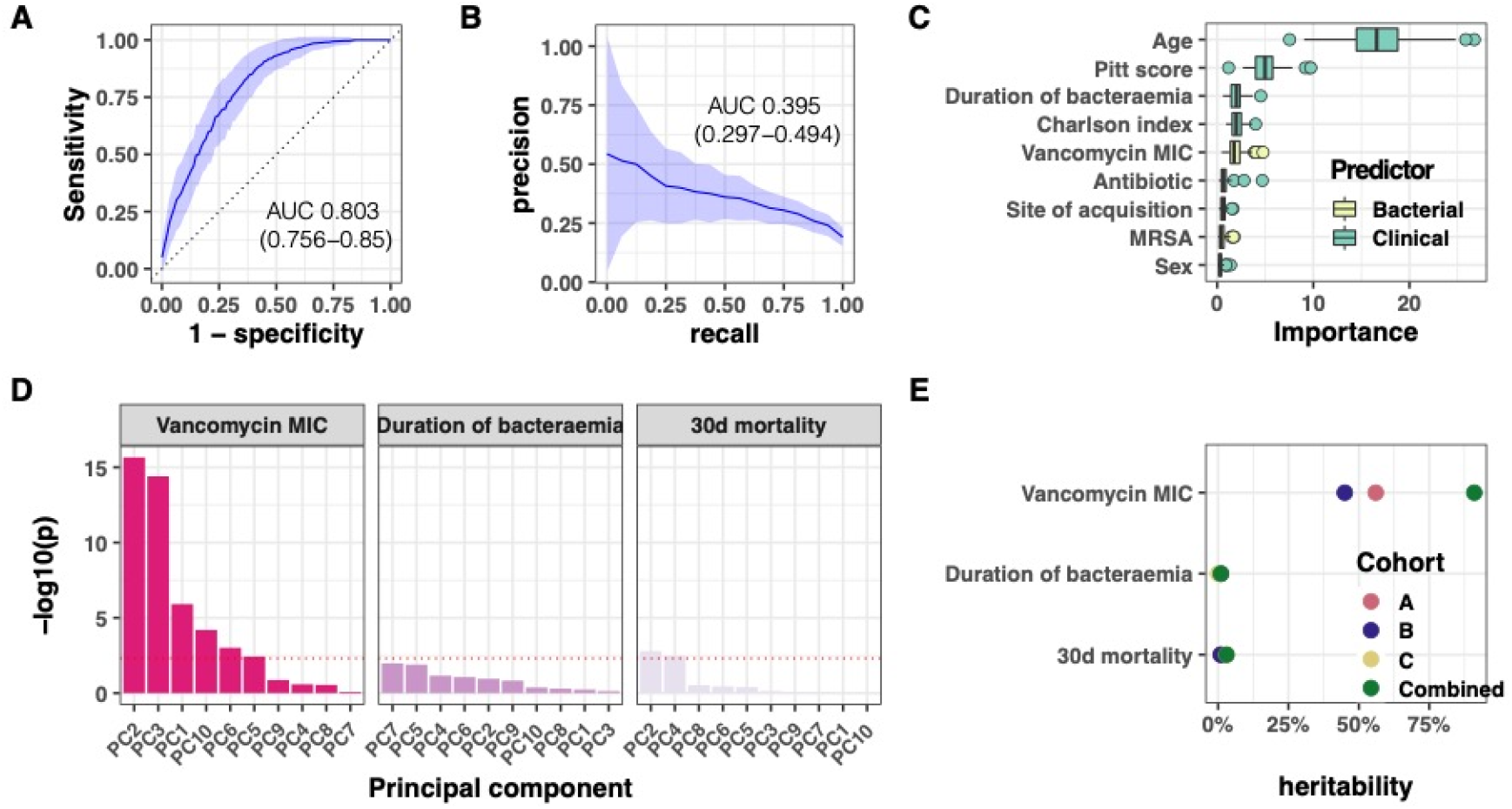
(A, B) Performance of the random forest model of 30-day mortality when using clinical and microbiological phenotypic data only. The model was trained and tested using cohort A data. The performance metrics (area under the curve [AUC]) in the test dataset were calculated from 100 replicate analyses. Panel A: receiver operator characteristic curve (ROC). Panel B: precision-recall curve (C) Variable importance plot (Gini impurity index) based on 100 replicate analyses. (D) Lineage effects for the vancomycin MIC, duration of bacteraemia and the SAB mortality outcomes for cohort A. The bars show the significance of the association between the ten first principal components (based on multidimensional scaling) and the outcome. (E) Narrow-sense heritability (SNP-heritability) of the GWAS outcomes of interest: vancomycin MIC, duration of bacteraemia (persistence) and SAB mortality.

**Figure 3.**
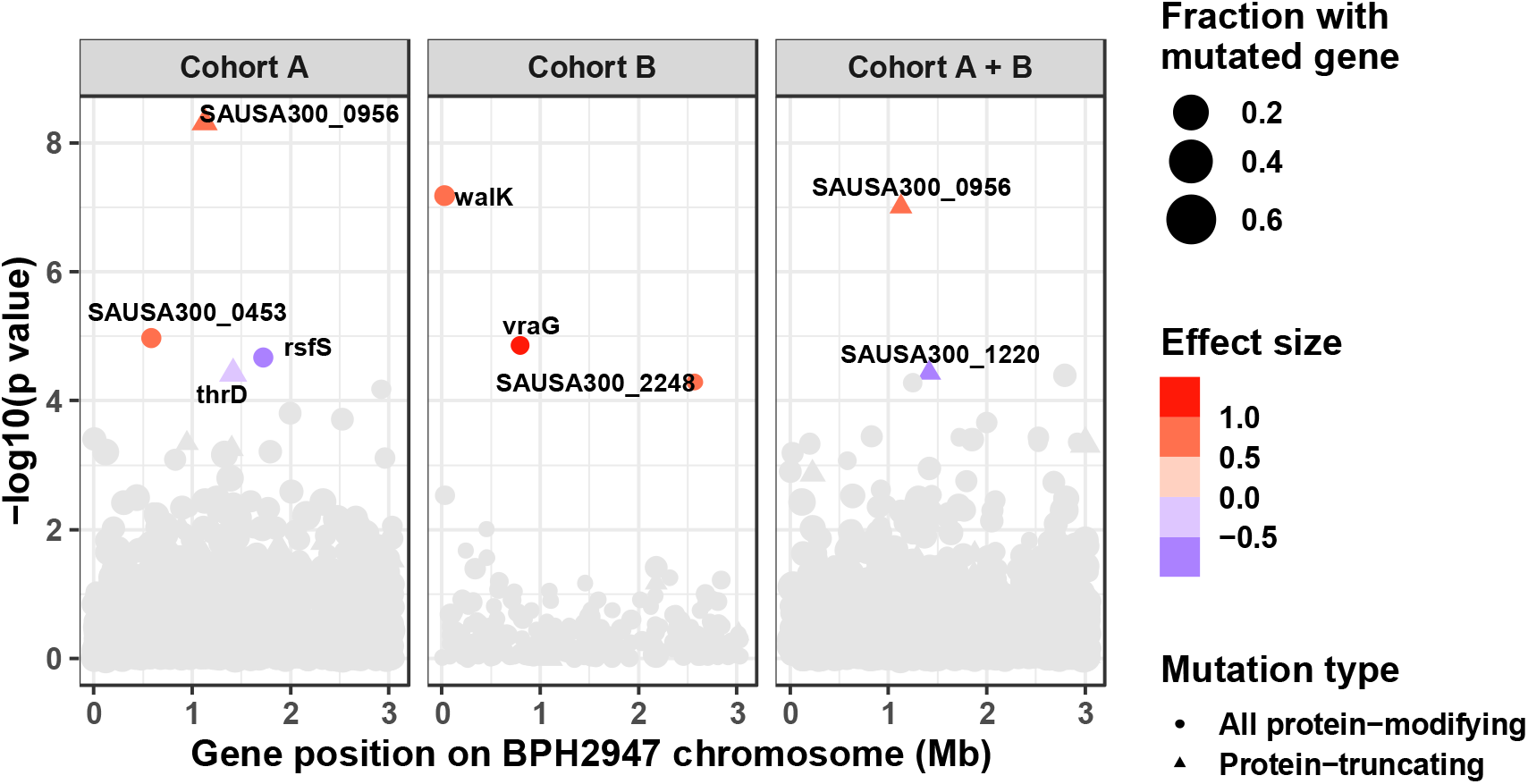
Gene-burden GWAS of vancomycin MIC in cohorts A, B and A and B combined. (no vancomycin E-test data were available for cohort C). The association was computed using a linear mixed model with vancomycin MIC as continuous outcome variable. All mutations with predicted impact on the protein sequence were aggregated by coding regions (circles: all protein-modifying, triangles: protein-truncating only). The size of the points represents the fractions of strains with a mutated genes and the colour indicated the effect size (impact on vancomycin MIC) in the linear regression.

To estimate the relative role of these predictors of mortality, we calculate variable importance (impurity index) based on 100 replicate analyses. While age and comorbidities were the most important features in the final model (as expected), we were surprised that vancomycin MIC was as important as comorbidities and more important than methicillin resistance (Figure 2C). This highlights the role of bacterial factors in SAB mortality and suggests that adaptive antibiotic resistance is more important than acquired antibiotic resistance genes.

To explore the relative role of bacterial genetic factors in SAB outcomes, we first considered whether there were “high-risk” clones or lineages that were associated with either of the three GWAS outcomes. To assess lineage effects in cohort A, we first performed a multi-dimensional scaling analysis (MDS) of a distance matrix generated by Mash, an approach that compares genetic similarity based on a subsample of 1,000 kmers (Ondov, Treangen et al. 2016). This allowed to include the whole genome content rather than just the core genome SNP matrix. We then extracted the top 10 components (85% of genetic variance explained) and assessed their correlation both with mortality and bacterial covariates. This correlation analysis showed robust lineage effects for vancomycin MIC, in particular PC2 and PC3 (p < 10^−15^, Figure 2D and Table S2), while no clear lineage effects were found for the clinical outcomes. As expected, there was a strong correlation between MDS axes and clonal complexes (CC) (Figure 1B and Figures S1 and S2). For example, the most prevalent clonal complex (CC 239) was positively correlated with PC2 and negatively correlated with PC1, while CC 22 was CC93 was strongly correlated with PC3.

We then calculated the narrow-sense heritability (h^2^) of clinical outcomes and bacterial phenotypes to determine what proportion of these parameters was a result of genetic factors. As expected, vancomycin MIC had a high heritability, while duration of bacteraemia and mortality had substantially lower values (Figure 2E and Table S1). These values are consistent bacterial GWAS of *S. aureus* infection vs. colonisation (Young, Wu et al. 2021) and *E. coli* bacteraemia mortality (Denamur, Condamine et al. 2022). We also sought to estimate the contribution of lineages to the phenotypic variance by extracting from the linear mixed model the variance attributed to the first 10 PC. We calculated that 87% of the vancomycin MIC variance could be explained by lineage effects, while lineage effects accounted for 12% of the mortality variance. This is consistent with the analysis of lineage-specific associations with vancomycin, with highly significant results for first three components (Figure 2D).

### Identification of loci associated with increased vancomycin MIC

We first analysed loci associated with vancomycin MIC (as a continuous variable) in two cohorts where vancomycin was determined using E-test. As E-test provides a more granular estimate of vancomycin MIC than broth microdilution (Holmes, Johnson et al. 2012), we expected to have an increased power to detect associations using a linear regression model. The MIC distribution was slightly shifted between the two cohorts with higher median MIC in cohort A than cohort B (1.5 mg/l vs 1mg/l, table 1, figure 1C), however, the shape of the MIC distribution was similar, suggesting that it would be possible to compare the results from the analyses performed in both cohorts separately.

Consistent with the heritability estimate, we identified several mutations that were significantly associated with vancomycin MIC, however the analysis was strongly biased by the population structure, as shown by the quantile-quantile plot (Figure S3). While this indicates inflation (despite correction for population structure in linear mixed model) and confirms the critical role of lineage effects in antibiotic resistance in *S. aureus*, close inspection of variants reaching genome-wide significance showed mutations in genes *mprF* (E269D, *mprF* G637S, *pbp2* A246T) known to be associated with antibiotic resistance in *S. aureus* (Machado, Seif et al. 2021), confirming the ability of this approach to identify resistance-associated loci (Table 2). In addition, we found an intergenic mutation 345 bp upstream of *mprF*, further supporting the importance of *mprF* in our datasets.

**Table 2.** Selection of loci significantly associated with vancomycin MIC. Loci are displayed if they have the lowest p value within the analysis or if they are located near or in known vancomycin resistance loci

| Reference position | Gene | Product | Mutation | P value (effect size) |  |  |
| --- | --- | --- | --- | --- | --- | --- |
|  |  |  |  | Cohort A | Cohort B | Combined analysis |
| 976503 | *sufU* | NifU family<br>SUF<br>system FeS<br>assembly<br>protein | V141I | $3.2 \times 10^{-12}$<br>(0.886) | | $7.5 \times 10^{-9}$<br>(0.877) |
| 1293856 | intergenic | Upstream of *def2-fmt-sun-rlmN-stp1-pknB* operon | | | | $6.51 \times 10^{-12}$<br>(0.779) |
| 2959009 | intergenic | Upstream of *aur* gene | | | $1.12 \times 10^{-4}$<br>(0.737) | |
| 1458535 | intergenic | Upstream of *mprF* | | $1.3 \times 10^{-11}$<br>(0.837) | | $5.52 \times 10^{-8}$<br>(0.827) |
| 1459663 | *mprF* | oxacillin<br>resistance-<br>related<br>FmtC<br>protein | E269D | $1.04 \times 10^{-7}$<br>(0.74) | | |
| 1460765 | *mprF* | oxacillin<br>resistance-<br>related<br>FmtC<br>protein | G637S | | | $7.12 \times 10^{-7}$ (-<br>0.479) |
| 1583057 | *pbp2* | penicillin<br>binding<br>protein 2 | A246T | | | $7.12 \times 10^{-7}$ (-<br>0.479) |

We also considered significant mutations outside known vancomycin resistance loci to explore whether our GWAS had the ability to identify new targets of vancomycin resistance. The most significant association in cohort A was found with the mutation V141I in the iron-sulfur cluster synthesis gene *sufU* (Roberts, Al-Tameemi et al. 2017) and with an intergenic mutation at position 1,293,856 of the reference genome *S. aureus* BPH2947 that was located within a promoter upstream of the predicted operon that contains the serine-threonine phosphatase *stp1*, which has been previously associated with vancomycin resistance (Cameron, Ward et al. 2012, Passalacqua, Satola et al. 2012). Only one locus reached genome-wide significance in cohort B: an intergenic mutation at position 2,959,009 within a predicted promoter upstream of the aureolysin gene *aur*.

Inflation of p values and incomplete correction for population structure indicated that GWAS at variant level was not sufficiently accurate to identify new targets. Moreover, it is known from within-host adaptive studies that convergence at mutational level is weak (Young, Wu et al. 2017). Therefore, we aggregated mutations at the gene level, an approach that has been used by others (Baines, Holt et al. 2015, Lees, Galardini et al. 2018, Saund and Snitkin 2020) and that may implicitly consider convergent evolution since different mutations are expected to arise independently across the phylogeny. Because the effect of non-synonymous substitutions could be both a gain or loss of functions, we analysed protein-modifying (all mutations excluding synonymous substitutions) and truncations separately and aggregated only rare mutations (allele fraction below 1%).

This analysis revealed 8 genes that reached genome-wide significance. Again, some of these genes (such as *walK and vraG* in cohort B) are known to be associated with vancomycin resistance but were not identified in the mutations analysis, highlighting the potential of the gene-burden test to increase the power of GWAS. A strong association was found with the SAUSA300_0956 locus, a GNAT-family acetyltransferase (Majorek, Osinski et al. 2017, Whaley, Radka et al. 2021). The function of the protein in *S. aureus* is not well understood, however, some indirect evidence may point to a role in cell-wall synthesis or turn-over. First, it is located between the autolysin *atl* and the methicillin resistance factor *fmtA (Qamar and Golemi-Kotra 2012)*. Second, an *E. coli* homolog of SAUSA300_0956 was linked to membrane biosynthesis (Whaley, Radka et al. 2021). Another interest association involved the SAUSA300_1220 locus. This gene (also known as *desR*) was recently found to be potentially associated with regulation of β-lactam resistance in MRSA (El-Halfawy, Czarny et al. 2020).

### Gene-burden GWAS identifies novel potential genetic signatures of persistence in *S. aureus* bacteraemia

We next assessed genetic correlates of duration of bacteraemia in cohort A and C. As expected, the distribution of this phenotype was skewed with 59 % of strains being associated with a duration of bacteraemia of 1 day or less (Figure 1D). Using an automated normalisation optimisation (based on Pearson P statistics), we found that none of the most common available strategies could improve the distribution of the variable (Figure S4).

No single site or mutation achieved genome-wide significance in accordance with a lower heritability and an expected polygenic signal associated with this complex trait (Dataset S3). However, we identified loci that were associated with duration of bacteraemia on the truncation burden test (gene-based and direction of change analysis). The strongest association in cohort A was found with protein truncations in *murQ*, an esterase that is part of the peptidoglycan recycling machinery (Borisova, Gaupp et al. 2016). Mapping of *murQ* truncations showed that they were independently acquired multiples times across the phylogeny, suggesting convergent evolution (Figure 4B).

**Figure 4.**
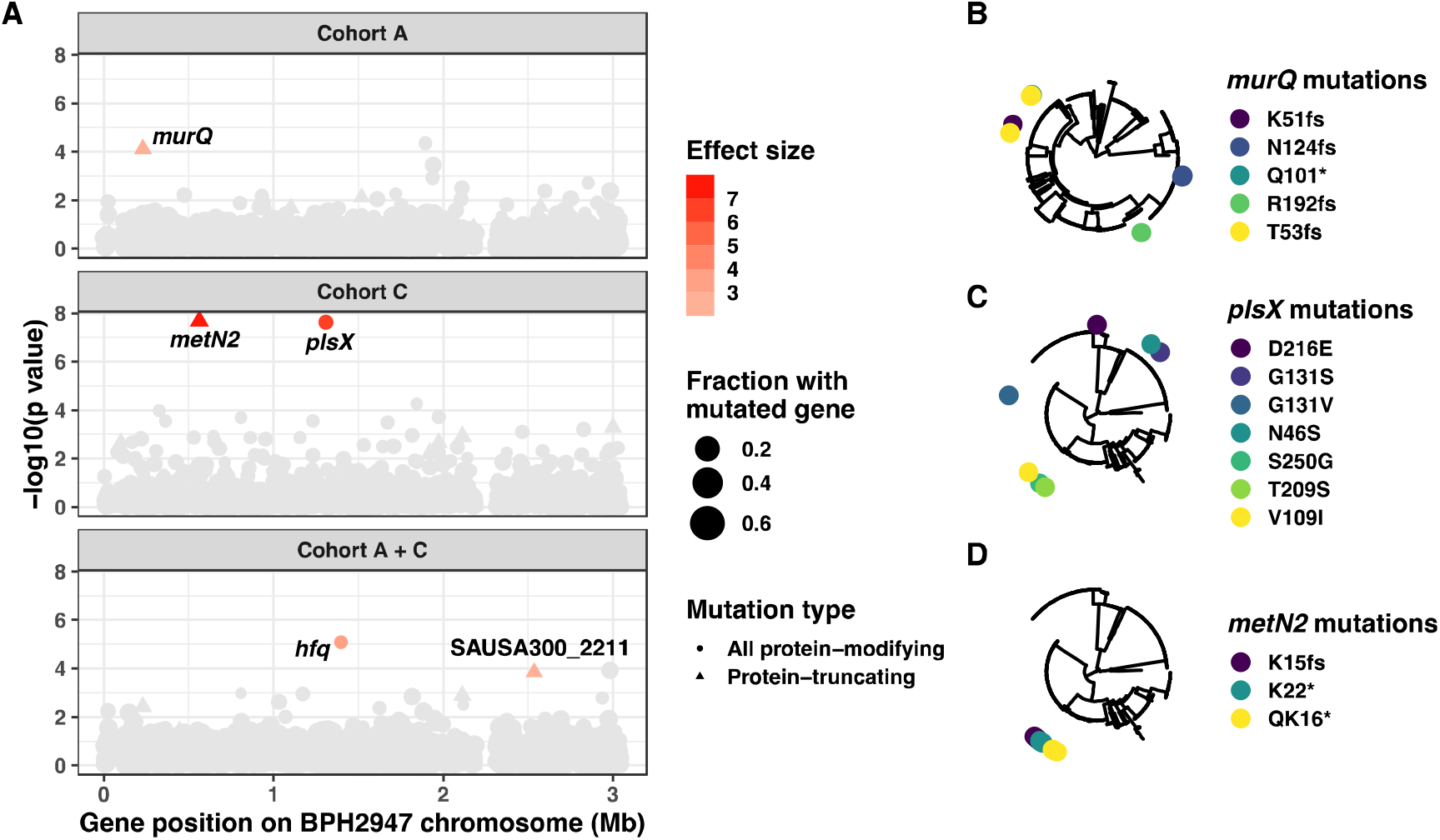
Gene-burden GWAS of duration of bacteraemia in cohorts A and C and A and C combined (duration of bacteraemia was not recorded for cohort B). (A) Manhattan plot showing the strength of the significance of mutated genes. The association was computed using a linear mixed model with duration of bacteraemia as continuous outcome variable. All mutations with predicted impact on the protein sequence were aggregated by coding regions (circles: all protein-modifying, triangles: protein-truncating only). The size of the points represents the fractions of strains with a mutated genes and the colour indicated the effect size (impact on duration of bacteraemia) in the linear regression. (B-D) Positions of *murQ* (B), *plsX* (C) and *metN2* (D) mutations on the phylogenetic tree.

**Figure 5.**
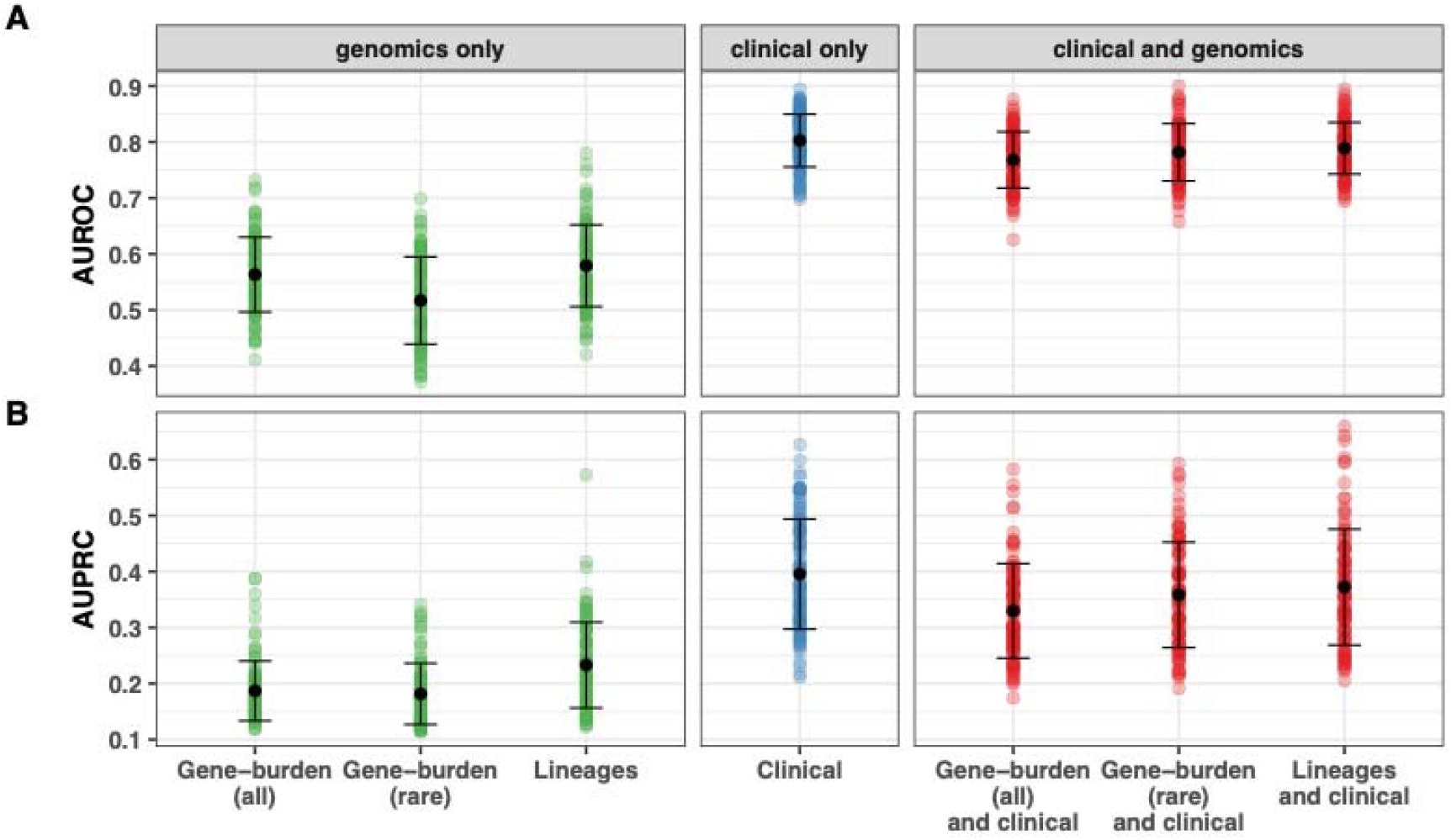
Predictive performance of seven random forest models of SAB mortality in cohort A. The data shown are the summary of 100 replicated random data-splits. Error bars represent mean and standard deviation. Panel A: area under the receiver operating characteristic curve (AUROC). Panel B: area under the precision-recall curve (AUPRC).

We found two significant associations in cohort C. These involved truncations in *metN2*, a methionine transporter, and protein-modifying mutations in *plsX*. The latter gene is a phosphate acyltransferase and is part of the phospholipid synthesis pathway (Whaley, Radka et al. 2021). It interacts with the fatty-acid kinase *fakA* which has been linked to virulence factors expression (Parsons, Broussard et al. 2014) and resistance to antimicrobial peptides (Li, Rigby et al. 2009).

### Bacterial genomic predictors don’t improve mortality prediction of *S. aureus* bacteraemia

We assessed whether there were any mutations or loci associated with SAB mortality in the three cohorts and the combined dataset. Only in cohort C could we detect a genome-wide significant association with rare mutations in four putative metabolic genes (SAUSA300_1088, *hxlA, glcU, ctaB*) (Figure S5).

This suggested that single locus effects don’t have a strong impact on mortality, or that this impact couldn’t be detected with datasets of our size. To investigate whether combination of mutations in multiple genetic loci could influence SAB mortality, we assessed the predictive power of bacterial genetic variation in our classification model of mortality. Since dimensionality can affect the performance of machine learning algorithms, we didn’t assess the single variants, but we rather built a genotype matrix with all mutated core genes with a score based on the predicted impact of the mutation (lowest score for no mutation, highest score for predicted protein truncations). The performance of this unfiltered gene-based approach was only slightly higher than chance alone (mean AUROC 0.56, mean AUPRC 0.17), and did not improve when restricting to rare mutations, using the same approach as with the gene-burden GWAS (mean AUROC 0.52, mean AUPRC 0.17). We also considered a model that used only the first ten principal components with similar predictive performance (mean AUROC 0.58, mean AUPRC 0.21). Furthermore, the performance of the model based on clinical predictors was not improved by adding genomic predictors (mean AUROC 0.77-0.78, mean AUPRC 0.31-0.35).

## DISCUSSION

We used a multifaceted strategy including bacterial GWAS and supervised machine learning to assess the role of bacterial genetic factors in clinical outcomes of *S. aureus* bacteraemia. We found that the bacterial genotype has limited impact on these outcomes, in particular mortality, as demonstrated by low heritability, poor predictive performance in our classification model of SAB and limited number of significant findings in the bacterial GWAS. Despite the weak genotype-outcomes association, we were able to identify a small number of candidate patho-adaptive loci associated with duration of bacteraemia, an important intermediate outcome of SAB that can reflect treatment response. In addition, our GWAS approach expanded the current picture of the the polygenic background of vancomycin resistance in clinical *S. aureus* with several new genetic loci implicated.

Identifying causative genomic correlates of bacterial phenotypes has been difficult even when dealing with phenotypes with expected strong heritability (like antibiotic resistance) (Chen and Shapiro 2021) but it is particularly challenging for clinical phenotypes such as invasiveness, treatment response or infection-related mortality (Lapp, Han et al. 2021, Young, Wu et al. 2021). These outcomes are expected to be multifactorial and the relative contribution of bacterial genetics can be extremely variable. Moreover, because the size of the datasets are usually small, negative findings might reflect an insufficient sample size, not an absence of contributing bacterial effects.

Among these multifactorial, clinical outcomes, invasiveness is thought to have a detectable bacterial molecular signature (Lees, Ferwerda et al. 2019). This is because even if bacterial invasion was exclusively determined by host and environmental factors (or by chance), some form of adaptive evolution would be expected as the result of the transition from the colonising niche to blood or human tissues (Culyba and Van Tyne 2021), as confirmed by within-host evolution studies where a proportion of invasive isolates carried molecular signatures of adaptation that were non present in colonising isolates (Lees, Kremer et al. 2017, Young, Wu et al. 2017). In accordance with this premise, two adequately sized bacterial GWAS of *S. pneumoniae* meningitis have identified point mutations in both pathogenesis and antibiotic resistance genes that were significantly associated with bacterial meningitis (Lees, Ferwerda et al. 2019, Li, Metcalf et al. 2019). Strikingly, in one study the association was replicated in an independent cohort (Li, Metcalf et al. 2019). In their study, Lees *et al* estimated the heritability of bacterial meningitis was estimated at a staggering 70% (Lees, Ferwerda et al. 2019). By contrast, heritability of *S. aureus* invasiveness was much lower (4%) in the GWAS study by Young et al (Young, Wu et al. 2021), where they identified a highly-significant association between mutations in the dihydrofolate reductase gene *dfrB* (associated with antibiotic resistance). This result is consistent with the association of penicillin-binding protein mutations with invasive *S. pneumoniae* infections and suggests an intriguing association between antibiotic resistance and invasiveness in bacteria, possibly due to the pleomorphic effects of antibiotic resistance and its impact on immune evasion and persistence (Gao, Cameron et al. 2013, Guerillot, Kostoulias et al. 2019).

The attempt of applying bacterial GWAS to infection outcomes (such as mortality) has been less successful than antibiotic resistance mechanisms. While Recker *et al*, were able to identify some molecular correlates of mortality from SAB (Recker, Laabei et al. 2017), a Danish study found no association between bacterial genetic determinants and occurrence of infectious endocarditis during SAB (Lilje, Rasmussen et al. 2017). Similarly, Lees *et al* found a stark contrast between the high heritability of the invasiveness phenotype and the null heritability of the meningitis outcome phenotype (Lees, Ferwerda et al. 2019). In a bacterial GWAS of a cohort of 912 patients with *Escherichia coli* bacteraemia, no bacterial genetic variant was associated with mortality, septic shock or intensive care unit admission. This correlated with very low heritability estimates for these outcomes (Denamur, Condamine et al. 2022).

Our heritability estimates and machine learning model confirmed the weak association between bacterial genotype and clinical outcomes. However, we hypothesised that it was possible to detect a role for bacterial variation based on the assumption that niche adaptation of invasive *S. aureus* promotes antibiotic resistance or tolerance. Building on this framework we performed a multi-cohort GWAS with a series of phenotypes with expected decreasing importance of bacterial factors (vancomycin MIC, duration of infection, treatment response and mortality). Specifically, we focused on the vancomycin MIC, because this phenotype was an important feature in our random forest model of mortality from SAB, in agreement with previous analyses of the cohort A (Holmes, Turnidge et al. 2011). In addition, vancomycin MIC is an example of adaptative phenotype that is relatively easy to measure and that is available for hundreds of clinical isolates. We then progressed to duration of bacteraemia, a key intermediary outcome of SAB (Kuehl, Morata et al. 2020) that is related to antibiotic efficacy and is often integrated into clinical management.

We first identified a key role of lineage effects, in particular the vancomycin MIC phenotype. Identifying lineage effects is important because they provide an estimate of the risk of false positive GWAS finding related to the confounding from the population structure (Earle, Wu et al. 2016). On the other hand, the presence of lineage effects might signify that there are high-risk clones for resistance or virulence phenotype, a feature that is important for pathogen surveillance and possibly for outcome prediction (Lam, Wick et al. 2021).

Previous GWAS of vancomycin MIC have shown the role of known determinants of vancomycin resistance including *walKR* (Baines, Holt et al. 2015), *rpoB* (Alam, Petit et al. 2014). Here, in a large GWAS of vancomycin MIC, we confirm the role of some of these established determinants of vancomycin resistance (*walK, mprF*), and provide evidence for new loci, the most promising being *desR*, a response regulator (Kim, Kim et al. 2016) that was recently singled out in a screening for targets of an antivirulence molecule that is able to reverse beta-lactam resistance in MRSA strains (El-Halfawy, Czarny et al. 2020). Our results confirm the potential for bacterial GWAS to assist the discovery of non-canonical antibiotic resistance loci (Farhat, Freschi et al. 2019).

Overall, only a few molecular markers achieve genome-wide significance in our GWAS of SAB mortality, indicating that larger sample sizes including thousands of sequences will be necessary to draw firm conclusion on the impact the bacterial genotype on mortality. Therefore, we considered duration of bacteraemia, an intermediate outcome that has strong association with mortality (Kuehl, Morata et al. 2020). We found that truncations in *murQ*, a gene degrades MurNAc-6p, an intermediate metabolite in peptidoglycan recycling (Borisova, Gaupp et al. 2016), were significantly associated with persistent SAB, potentially suggesting a role in antibiotic resistance or tolerance. Two candidate loci of persistent bacteraemia were the methionine biosynthesis locus *metN2* and the phospholipid biosynthesis gene *plsX*. MetN2 is an ABC-transporter; a potential clue to its role in bacteraemia comes from a study where expression of *metN2* was strongly reduced in *S. aureus* mutants that were resistant to antimicrobial peptides (Kubicek-Sutherland, Lofton et al. 2017). Interestingly, *plsX* has also a potential association with antimicrobial peptide resistance: it is closely connected with the fatty acid kinase *fakA* (Parsons, Broussard et al. 2014), which was found to be linked to resistance to the antimicrobial peptide dermcidin in a transposon mutant screen (Li, Rigby et al. 2009).

There is ongoing uncertainty around the role of bacterial genetic factors in human infections. Here we show that despite a likely preponderant role for host and treatment factors in shaping infection outcomes, it is also possible to disentangle the multifactorial networks underlying these infections and detect specific bacterial genetic loci that help drive particular clinical outcomes. While these single loci likely have a low global predictive value, they might be more relevant in specific, high-risk settings (e.g. persistent or recurrent infection) and might act in concert with other bacterial variants, host genetic variants and treatment factors. In addition, the genetic loci that were enriched in our analysis might represent new clinically relevant molecular targets of antibiotic resistance and tolerance and important candidates for functional validation.

## METHODS

### Study cohorts

Cohort A was assembled from two multicentre observational studies of SAB that were performed to assess the impact of vancomycin MIC on clinical outcomes (Giulieri, Baines et al. 2018). The vancomycin substudy of the Australian and New Zealand Cooperative on Outcome in Staphylococcal Sepsis (ANZCOSS) study included 532 cases of SAB from a larger retrospective study of SAB between 2007-2008 (Turnidge, Kotsanas et al. 2009). The selection of episodes for the substudy has been described previously (Holmes, Turnidge et al. 2011). Briefly, to ensure a balanced proportion of cases, each SAB case treated with vancomycin was matched with the next available case treated with flucloxacillin at each participating centre. The Vancomycin Efficacy in Staphylococcal Sepsis in Australasia (VANESSA) study included prospectively 226 patients with SAB between 2012-2013 (Holmes, Robinson et al. 2018). Data collected within cohort A included patient demographics, site of acquisition, comorbidities, clinical syndrome, antibiotic treatment, 30-day mortality and duration of bacteraemia. Baseline isolates were collected for all patients, while supplementary isolates (colonising isolates from nasal swabs and serial blood isolates) were obtained when available. Cohort A had Cohort B included 296 episodes of MRSA bacteraemia due to sequence type 239 that were collected at a single centre in Australia between 1996-2008 to assess the impact of vancomycin MIC and vancomycin heteroresistance on 30 day mortality (van Hal, Jones et al. 2011).

Cohort C included data and sequences from the CAMERA2 trial (Combination Antibiotics for Methicillin Resistant *Staphylococcus aureus*) that was performed between 2015 and 2018 in Australia, New Zealand, Singapore and Israel, and randomised participants with MRSA bacteraemia to either monotherapy with vancomycin or daptomycin or combination therapy with vancomycin or daptomycin plus an antistaphylococcal beta-lactam (flucloxacillin, cloxacillin or cefazolin) (Tong, Lye et al. 2020). The primary outcome was a composite endpoint ascertained 90 days after randomisation and included mortality, persistent bacteraemia after 5 days, microbiological relapse (positive blood culture after a preceding negative culture), and microbiological treatment failure (positive sterile site culture 14 or more days after randomisation). In addition, the trial collected information on the day of death, duration of bacteraemia and day of relapse. Relevant clinical data assessed were comorbidites, severity scores, primary source and secondary foci, source control. Details of the antibiotic treatment were collected up to day 90.

### Phenotypic testing

Bacterial isolates were stored in glycerol broth at -80C, after confirmation of *S. aureus* using coagulase and DNase tests or Matrix-assisted laser desorption-ionisation time-of-flight (MALDI-TOF). Vancomycin minimum inhibitory concentration (MIC) was determined by E-test (bioMerieux) following manufacturer’s instructions. An inoculum of 0.5 McFarland was used. Vancomycin E-test was only performed for cohort A and B strains.

### Machine learning model to select predictors of mortality/treatment response

Using data from cohort A, we fitted a classification model of 30-day mortality of SAB to assess the predictive performance of non-genomic features including demographic and clinical data (age, Pitt bacteraemia score, Charlson comorbidity index, site of acquisition, gender), microbiological phenotypes (methicillin resistance, vancomycin MIC) and treatment response variables (duration of bacteraemia). The R package tidymodels was used to build a random forest model of SAB mortality. The model was trained on 80% of the data using stratified 10-fold cross-validation and hyperparameter tuning to optimise the area under the receiver operator characteristics curve (AUROC). Tuning of the random forest hyperparameters “mtry” (number of randomly selected predictors) and “min_n” (minimal node size) was done using a 5×5 grid, with range of hyperparameters determined empirically based on initial tuning using 20 automatically selected candidate combinations. Categorical predictors were converted into binary variables an all numeric predictors were centered and scaled. Zero variance predictors were removed. Downsampling was used to compensate for class inbalance (mortality rate 15%). The model was fitted on 100 replicated data-splits to generate estimates of the performance and variable importance as in (Lapp, Han et al. 2021).

### Whole-genome sequencing

Bacterial strains were subcultured twice from -80C glycerol stock. The Janus® automated workstation (PerkinElmer) or manual extraction kits (Invitrogen PureLink genomic DNA kit or the Sigma GenElute kit) were used to extract genomic DNA from single colonies. DNA concentrations were normalised to 0.2 ng/ul for library preparation using Nextera® XT DNA (Illumina). Whole-genome sequencing was performed on the Illumina MiSeq and NextSeq platforms. Reads were assembled using Shovill v1.1.0 (https://github.com/tseemann/shovill), a pipeline that streamlines and optimises Spades (58). To assess the quality of the sequences, mean read depth, assembly metrics and the percentage of *S. aureus* reads using Kraken 2 v2.1.1 (60), were calculated. Reads were discarded if read depth was < 20, the fraction of *S. aureus* reads was < 80% or the size of the assembly was > 3.5 Mb or < 2.5 Mb. We used Mash v2.3 to generate a distance matrix from the Shovill assemblies using 1,000 sketches.

### Variant calling

Reads were mapped to reference genome *S. aureus* BPH2947 (accession GCF_900620245.1) using Snippy v4.6.0 (https://github.com/tseemann/snippy). For each dataset, a core genome alignment including all positions with at least 90% non-gaps was constructed using Snippy-core v4.6.0, Goalign v0.3.4 (Lemoine and Gascuel 2021) and SNP-sites v2.5.1 (Page, Taylor et al. 2016). The alignment was then used to infer a maximum-likelihood phylogenetic tree with IQ-TREE v2.0.3 (Minh, Schmidt et al. 2020) under a GTR+G4 model. Support was calculated using the ultrafast support feature and SH-aLRT test. For each variant at positions included in the core genome alignment, we computed the number of independent acquisitions across the phylogeny using HomoplasyFinder (Crispell, Balaz et al. 2019).

### Preparation of the bacterial genotype files

For each dataset, variants at core genome positions (minimum 90% of sequences with a coverage of at least 10 reads) were merged in a single variant call format (VCF) file using BCFtools v 1.10.2 (Danecek, Bonfield et al. 2021), without merging multiallelic records. A catalogue of mutations with predicted impact on protein function was generated by masking synonymous variants were masked using the command bcftools view -e ‘ANN[0] ∼ “[^&]synonymous”‘. Mutations with predicted loss of function (LOF) were extracted using the command bcftools view -i ‘ANN[0] ∼ “frameshift_variant” | ANN[0] ∼ “stop_gained” | ANN[0] ∼ “stop_lost” | ANN[0] ∼ “start_lost”‘.

### Variant annotation

In addition to the variant annotation based on BPH2947 generated by Snippy, we used Blast v 2.10.1 to determine homologs in reference genome FPR3757 that were linked to the AureoWiki (Fuchs, Mehlan et al. 2018) and MicrobesOnline (Dehal, Joachimiak et al. 2010) databases. Blast hits were kept if alignment coverage was above 50 % and identity was above 90 %. When multiple hits were available, the hits with highest identity were selected.

### Bacterial GWAS design

We ran bacterial GWAS to test the association between mutations with predicted impact on protein function (filtered as described above) with the phenotypes of interest, as continuous (vancomycin MIC, duration of bacteraemia) or binary traits (30 day mortality). For clinical outcomes, we analysed one sample per episode. For vancomycin MIC GWAS, we included more than one sample per episode when they were available. For duration of bacteraemia, potential transformations to achieve a normal distribution were assessed using the bestNormalize package in R (Peterson and Cavanaugh 2020).

### Bacterial GWAS pipeline

We used the factored spectrally transformed linear mixed models (FaST-LMM) as implemented in Pyseer v 1.1.2 (Lees, Galardini et al. 2018), which allows to correct for population structure by adding a kinship matrix as a random effect. The kinship matrix was computed from a VCF file including all core-genome variants using the Gemma, version 0.98.1 (Zhou and Stephens 2012). We assessed phenotype associations both with individual mutations and with protein-modifying and loss-of-function aggregated in genes, operons and gene ontology categories as implemented with the ‘--burden’ option in Pyseer. For the mutation GWAS, we excluded synonymous substitutions and mutations that were acquired only once across the phylogeny (based on homoplasy counts obtained using HomoplasyFinder (Crispell, Balaz et al. 2019)). For the gene-burden GWAS, only rare variants (i.e. those with a minim allele fraction below 1%) were considered.

### Lineage effects and heritability

Heritability (SNP-heritability, h^2^) - the proportion of phenotypic variance that is correlated with genotypic variance - was estimated by Pyseer from the linear mixed model using the kinship matrix computed in Gemma (see above). Lineages of cohort A isolates were inferred from the Mash distance matrix using the ‘cmdscale’ function in R. The association between the first 10 components and the bacterial GWAS outcomes was computed by running Pyseer with the ‘—lineage’ option. Correlation between the 10 first components and internal nodes of the tree or major clonal complexes was calculated as performed in the R package bugwas https://github.com/sgearle/bugwas/blob/master/bugwas/R/BUGWAS_functions.R#L962) (Earle, Wu et al. 2016).

## Data Availability

The raw reads of the strains included in this analysis are available in the European Nucleotide Archive under Bioproject accession numbers PRJEB27932 (cohort A), PRJEB50796 (cohort B) and PRJEB2561 (cohort C).

## Ethics approval

Human research committee approval was obtained at each site for the three cohorts. Written informed consent was collected for the prospective component of cohort A (VANESSA) and for cohort C (CAMERA2).

## Code availability

The code for the preparation of bacterial genotype files and for running Pyseer is available at https://github.com/stefanogg/CLOGEN.

