## Supplementary materials for "A statistical genomics framework to trace bacterial genomic predictors of clinical outcomes in *Staphylococcus aureus* bacteraemia"

**This PDF file includes:**

Tables S1 and S2

Figs. S1 to S5

Legends for Datasets S1 to S5

**Other Supplementary Materials for this manuscript include the following:**

Datasets S1 to S5

Code used to train and test the random forest models of *S. aureus* bacteraemia mortality

**Table S1.**

Heritability estimates for the GWAS outcomes vancomycin MIC, duration of bacteraemia and 30 day mortality. Heritability was calculated in Pyseer using a kinship matrix generated by Gemma with all core genome mutations.

| Phenotype | Cohort | Heritability |
| --- | --- | --- |
| Vancomycin MIC | A | 0.56 |
| Vancomycin MIC | B | 0.45 |
| Vancomycin MIC | Combined | 0.91 |
| Duration of bacteraemia | A | 0.01 |
| Duration of bacteraemia | C | 0 |
| Duration of bacteraemia | Combined | 0.01 |
| 30d mortality | A | 0.01 |
| 30d mortality | B | 0.01 |
| 30d mortality | C | 0.03 |
| 30d mortality | Combined | 0.03 |

**Table S2.**

Lineage effects in cohort A for the GWAS outcomes vancomycin MIC, duration of bacteraemia and 30 day mortality. Lineage effects were calculated in Pyseer using the first ten principal components obtained by multidimensional scaling of a distance matrix calculated with Mash.

| Phenotype | PC | Wald statistics | P value |
| --- | --- | --- | --- |
| Vancomycin MIC | MDS2 | 9.352 | < 2.22E-16 |
|  | MDS3 | 7.855 | 4.00E-15 |
|  | MDS1 | 4.852 | 1.23E-06 |
|  | MDS10 | 3.997 | 6.40E-05 |
|  | MDS6 | 3.296 | 9.82E-04 |
|  | MDS5 | 2.896 | 3.78E-03 |
|  | MDS9 | 1.492 | 1.36E-01 |
|  | MDS4 | 1.125 | 2.60E-01 |
|  | MDS8 | 1.067 | 2.86E-01 |
|  | MDS7 | 0.213 | 8.31E-01 |
| Duration of bacteraemia | MDS7 | 2.557 | 1.06E-02 |
|  | MDS5 | 2.486 | 1.29E-02 |
|  | MDS4 | 1.815 | 6.95E-02 |
|  | MDS6 | 1.715 | 8.64E-02 |
|  | MDS2 | 1.587 | 1.13E-01 |
|  | MDS9 | 1.444 | 1.49E-01 |
|  | MDS10 | 0.798 | 4.25E-01 |
|  | MDS8 | 0.662 | 5.08E-01 |
|  | MDS1 | 0.554 | 5.80E-01 |
|  | MDS3 | 0.330 | 7.41E-01 |
| 30d mortality | MDS2 | 3.152 | 1.62E-03 |
|  | MDS4 | 2.939 | 3.29E-03 |
|  | MDS8 | 1.031 | 3.03E-01 |
|  | MDS6 | 0.918 | 3.58E-01 |
|  | MDS5 | 0.833 | 4.05E-01 |
|  | MDS3 | 0.345 | 7.30E-01 |
|  | MDS9 | 0.194 | 8.46E-01 |
|  | MDS7 | 0.184 | 8.54E-01 |
|  | MDS1 | 0.090 | 9.28E-01 |
|  | MDS10 | 0.083 | 9.34E-01 |

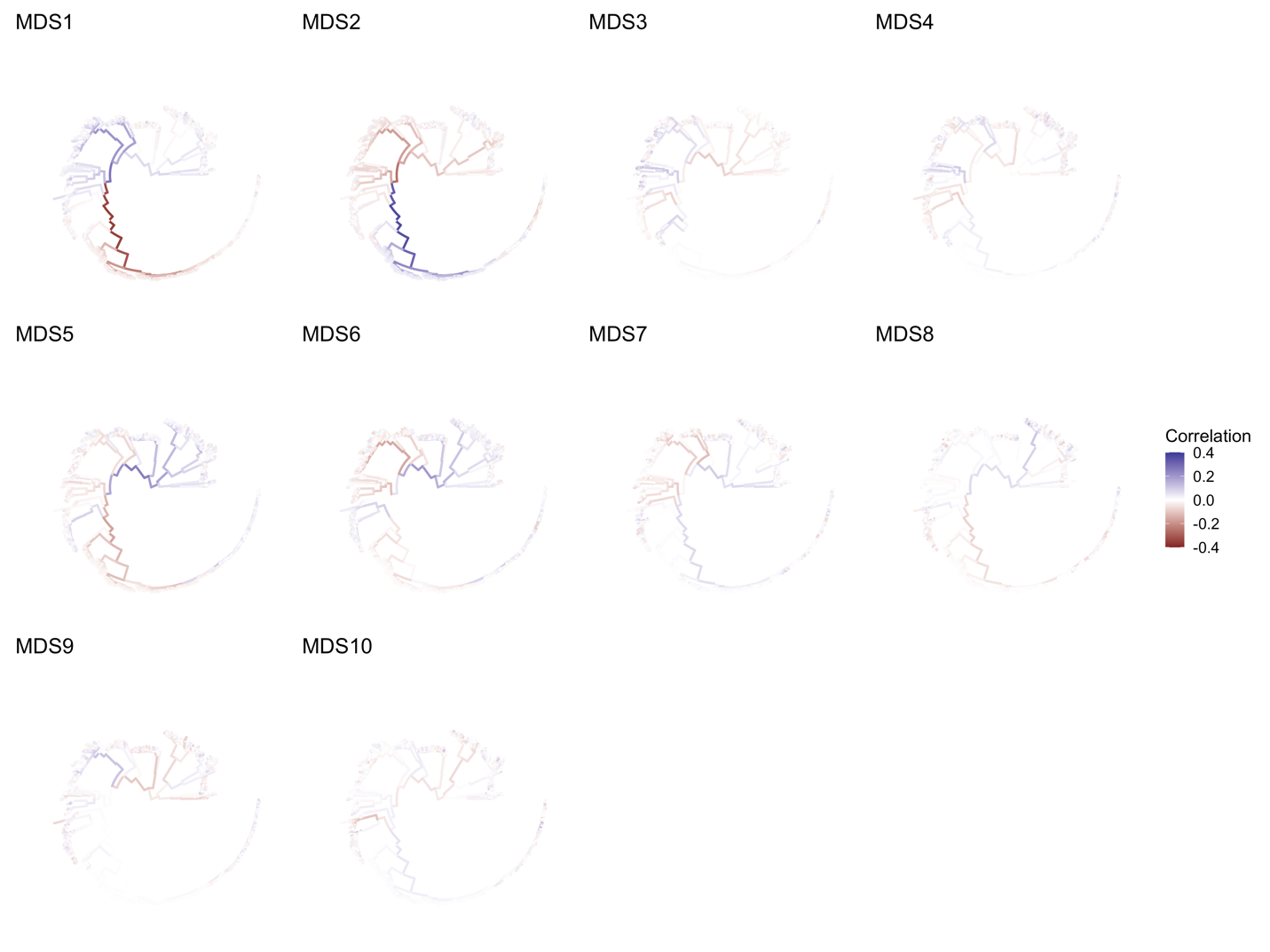

Figure S1.

Mapping on the phylogenetic tree of the 10 first principal components (generated by multidimensional scaling of the distance matrix obtained using Mash). Each component was correlated with the internal nodes.

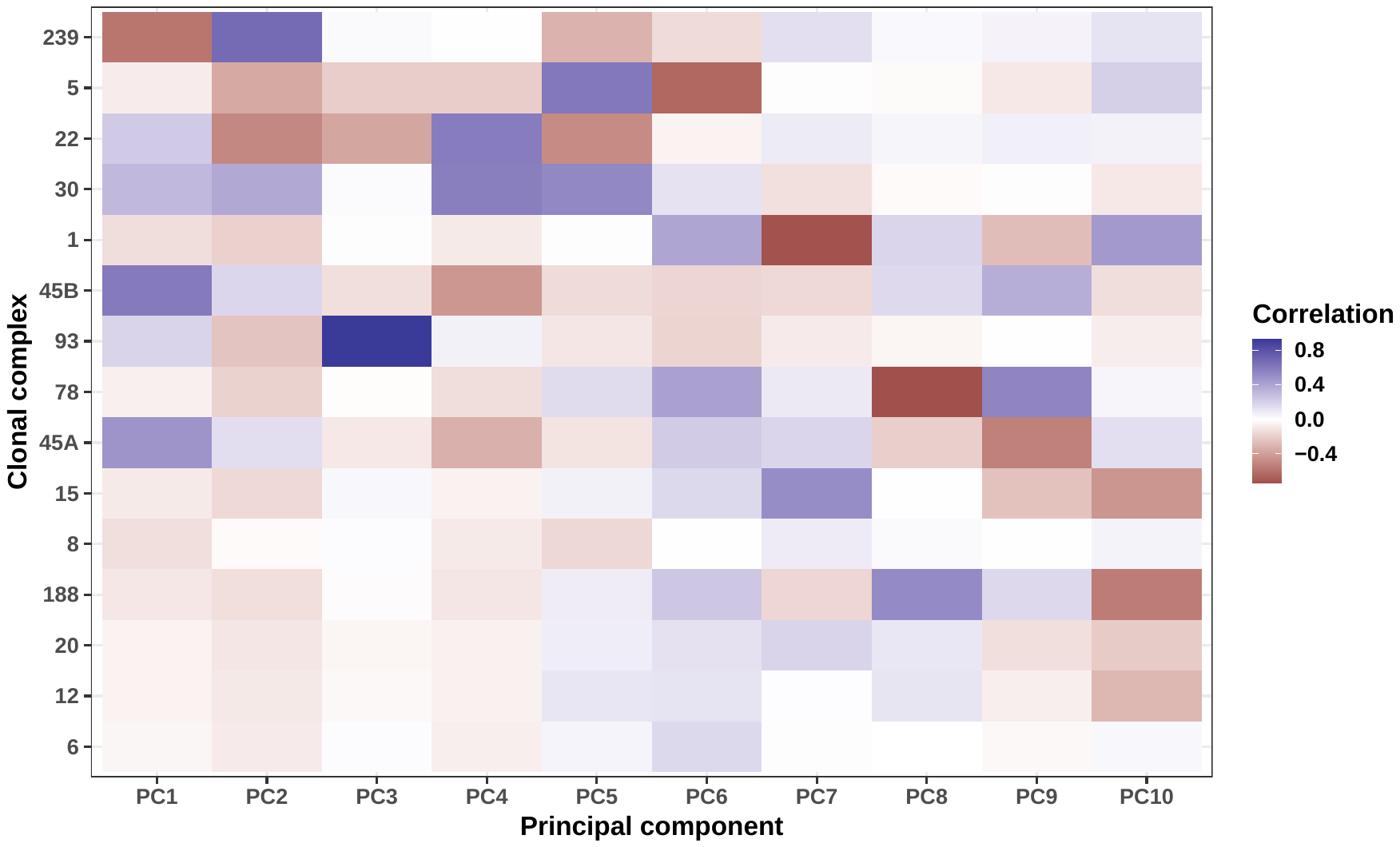

Figure S2.

Heatmap representing correlations between the top 15 clonal complexes and the first ten principal components generated by multidimensional scaling of the distance matrix obtained using Mash. The clonal complexes are ordered by the number of isolates, with CC239 being the largest clonal complex in the combined dataset.

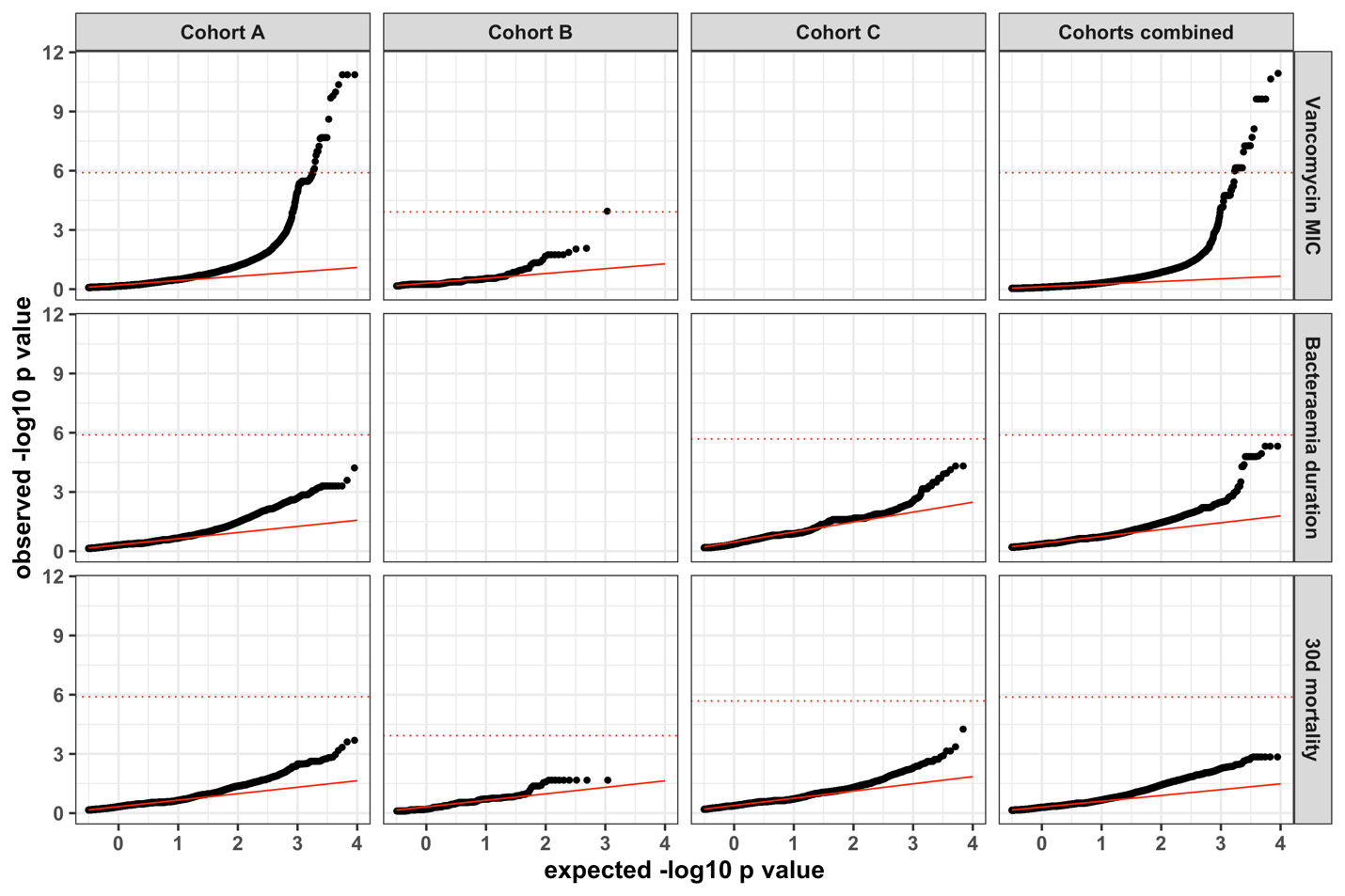

Figure S3.

Quantile-quantile plots of core genome mutations GWAS for all cohorts and cohort combinations (row facets) and for three outcomes vancomycin MIC, bacteraemia duration and 30-day mortality (column facets). The dotted red lines represent the Bonferroni-corrected significance threshold and the continuous read line shows the slope and intercept of the theoretical sample distribution.

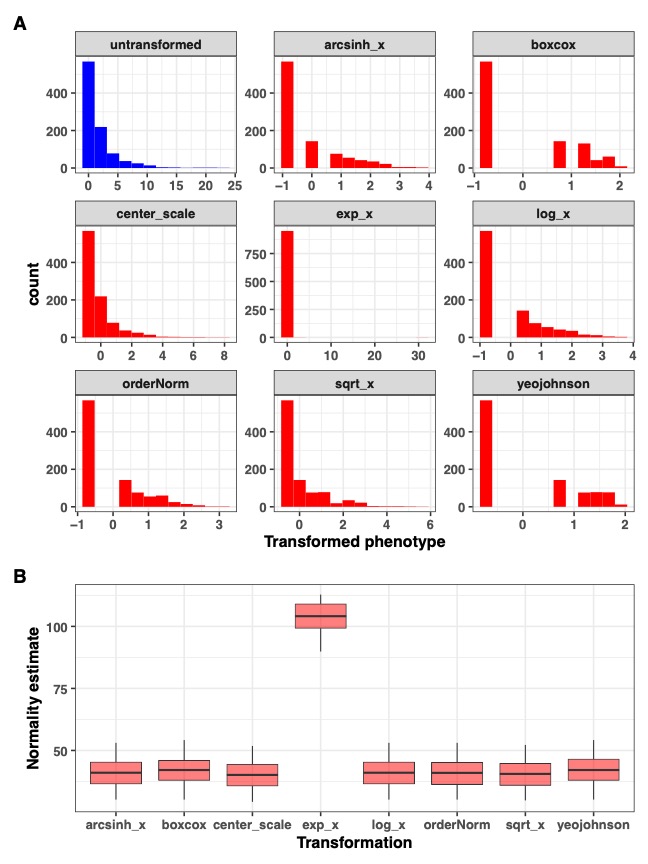

Figure S4.

Impact of eight automated normalisation algorithms on the bacteraemia duration in cohort A and C combined. The transformations were obtained with the R package bestNormalize. Panel A: histograms with distribution of the untransformed (blue) and transformed (red) variable. Panel B: out-of-sample estimate of normality statistics (Pearson’s P divided by its degree of freedom) for each transformation obtained using 10-fold cross-validation and 5 repeats. Transformations with the lower values are closer to normality. Note that the center scale transformation is equivalent to untransformed.

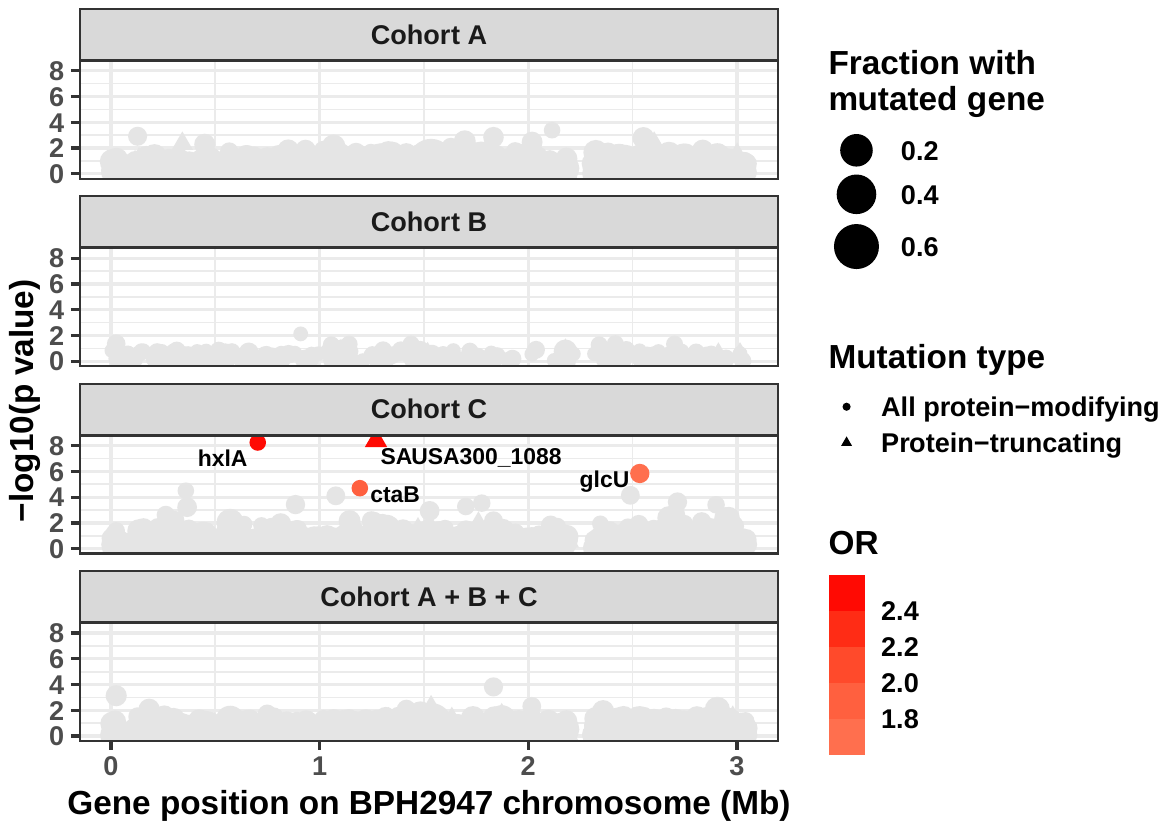

Figure S5.

Gene-burden GWAS of 30 day mortality in cohorts A, B, C and A+B+C combined. The Manhattan plot shows the strength of the significance of mutated genes. The association was computed using a linear mixed model with mortality as binary variable. All mutations with predicted impact on the protein sequence were aggregated by coding regions (circles: all protein-modifying, triangles: protein-truncating only). The size of the points represents the fractions of strains with a mutated genes and the colour indicated the effect size (odds ratio for mortality) in the logistic regression.

Dataset S1. (Dataset_S1.xlsx)

List of strains.

Dataset S2. (Dataset_S2.xlsx)

Output of vancomycin MIC GWAS.

Dataset S3. (Dataset_S3.xlsx)

Output of duration of bacteraemia GWAS.

Dataset S4 (Dataset_S4.xlsx)

Output of 30 day mortality GWAS.

Dataset S5 (Dataset_S5.xlsx)

Predictive performance and variable importance of the random forest models of *S. aureus* bacteraemia mortality.
